# Role of Inflammation in Depression and Anxiety: Tests for Disorder Specificity, Linearity and Potential Causality of Association in the UK Biobank

**DOI:** 10.1101/2021.02.02.21250987

**Authors:** Zheng Ye, Nils Kappelmann, Sylvain Moser, George Davey Smith, Stephen Burgess, Peter B. Jones, Golam M. Khandaker

**Affiliations:** Department of Psychiatry, University of Cambridge, Cambridge, United Kingdom; Department of Research in Translational Psychiatry, Max-Planck-Institute of Psychiatry, Munich, Germany; International Max Planck Research School for Translational Psychiatry (IMPRS-TP), Munich, Germany; MRC Integrative Epidemiology Unit, University of Bristol, Bristol, UK; Population Health Sciences, Bristol Medical School, University of Bristol, Bristol, UK; MRC Biostatistics Unit, University of Cambridge, Cambridge, UK; Cardiovascular Epidemiology Unit, Department of Public Health and Primary Care, University of Cambridge, Cambridge, UK; Cambridgeshire and Peterborough NHS Foundation Trust, Cambridge, UK; Centre for Academic Mental Health, Population Health Sciences, Bristol Medical School, University of Bristol, Bristol, UK

**Keywords:** Depression, Anxiety, Inflammation, CRP, IL-6, Mendelian randomisation

## Abstract

**Background:** Concentrations of C-reactive protein (CRP), interleukin 6 (IL-6) and other inflammatory markers are elevated in people with depression and anxiety compared to controls, but evidence for disorder-specificity, linearity and potential causality is sparse.

**Methods:** Using data from up to 144,890 UK Biobank cohort participants, we tested associations of circulating CRP concentrations with depression and anxiety symptom scores and probable diagnosis, including tests for linearity, disorder-specificity and sex difference. We examined potential causality using 1-sample and 2-sample Mendelian randomisation (MR) analyses testing associations of genetically-predicted CRP concentration and IL-6 activity with depression and anxiety.

**Findings:** CRP concentration was associated with depressive and anxiety symptom scores and with probable diagnoses of depression and generalised anxiety disorder (GAD) in a dose-response fashion. These associations were stronger for depression than for anxiety, and for women than for men although less consistently. MR analyses provided consistent results suggesting that genetically predicted higher IL-6 activity was associated with increased risk for depressive symptoms, while genetically-predicted higher CRP concentration was associated with decreased risks of depressive and anxiety symptoms.

**Interpretation:** Altered activity of the IL-6/IL-6R pathway could be causally linked to depression. The field now requires experimental studies of IL-6 modulation in humans and animal models to further examine causality, mechanisms and treatment potential. Such studies are also needed to elucidate mechanisms for divergent associations of genetically-predicted higher IL-6 activity (risk increasing) and higher CRP concentrations (protective) with depression/anxiety.

**Funding:** MQ (MQDS17/40); Wellcome Trust (201486/Z/16/Z).

## INTRODUCTION

Innate immune dysfunction represents a putative mechanism for depression and other psychiatric disorders opening up the possibility of new treatment approaches distinct from current monoaminergic drugs.^1,2^ In depression, for instance, there is evidence of low-grade systemic inflammation as indexed by elevated concentrations of C-reactive protein (CRP >3mg/L) in 21–34% of patients,^3^ along with increased concentrations of interleukin-6 (IL-6) and other inflammatory cytokines in blood and in cerebrospinal fluid (CSF).^4,5^ A number of randomised controlled trials (RCTs) are now testing the effects of anti-inflammatory drugs in patients with depression (e.g., Khandaker *et al*.^6^, NCT02473289, NCT02362529). However, there are key outstanding questions, particularly regarding specificity and causality of association, that require addressing for a clearer understanding of the potential role of inflammation in illness pathogenesis and to inform future clinical trials.

Depression overlaps with anxiety both genetically and clinically. Anxiety symptoms now form part of the diagnostic criteria for major depressive disorder (MDD) in the diagnostic and statistical manual of mental disorders 5^th^ edition (DSM-5).^7^ However, to our knowledge no studies have tested whether inflammation is a common or specific risk factor for depression and anxiety. This is an important issue as it may help to identify potentially unique or shared mechanisms for psychiatric disorders that commonly co-occur.

Regarding causality, longitudinal studies have reported evidence for a temporal association between elevated CRP and IL-6 concentrations at baseline and risk of depression subsequently,^8,9^ but residual confounding still remains a possibility. Mendelian randomization (MR) is an epidemiological approach that uses genetic variants as instruments to untangle the problem of unmeasured confounding as genetic variants are randomly inherited from parents to offspring and fixed at conception.^10^ Therefore, if genetically-predicted values of a risk factor are associated with a disease outcome, then it is likely the association between the risk factor and outcome has a causal basis.

Existing MR studies have provided mixed evidence on the association of inflammation with different psychiatric disorders. Hartwig *et al*. reported potential protective effects of elevated CRP for schizophrenia,^11^ contrasting with findings from observational studies.^12,13^ For depression, one study did not find evidence for a potential causal role of inflammation,^14^ while more recent studies reported potential causal roles for increased IL-6 and CRP serum concentrations in depression^15^ and for increased IL-6 activity for suicidality specifically.^16^

While these findings may indicate disorder-specificity, further research is required to enable definite conclusions regarding causality of association. Furthermore, to our knowledge, MR studies of inflammation and anxiety are lacking.

We have used data from up to 144,890 individuals from the UK Biobank study, a large general population-based cohort, to test associations of circulating CRP concentrations with depression and anxiety. As outcomes, we have used symptom scores and categorical probable diagnosis in the total sample and in men and women separately to assess potential sex difference, strength and reproducibility of association. We have examined evidence for dose-response by testing linearity of association. We have examined specificity of association by testing whether the association of CRP with depression and anxiety is stronger for one outcome than the other, or is similar between outcomes. Furthermore, we have carried out MR analysis in the full sample, and in men and women separately, to test whether associations of CRP and IL-6 with depression and anxiety are consistent with potential causal roles for these biomarkers in these conditions.

## METHODS

### Study population

The UK Biobank is a population-based cohort with a range of phenotyping assessments, biochemical assays and genome-wide genotyping from over 500,000 UK residents aged 40-69 years at baseline, recruited between 2006 and 2010 from 22 assessment centres throughout the UK.^17^ Our primary outcomes were depressive and anxiety symptoms that were assessed online as part of a follow-up mental health survey completed by up to 157,115 individuals.^18^ The current study used available data from the maximum number of UK Biobank participants for each analysis (N up to 144,890). The UK Biobank study was subject to ethics committee approval and participants gave their informed consent prior to participation; see details in Supplementary Methods.

### Exposure

Serum high-sensitivity CRP concentrations were measured by immunoturbidimetric assay on a Beckman Coulter AU5800. Minimum detection limit was 0.08 mg/L. CRP values in the entire sample (n=486,424) ranged from 0.08 to 79.96 mg/L; mean=2.60 (SD=4.36) mg/L. The distribution of CRP concentrations for this study (n=146,954) was divided into quintiles or deciles, which were used as categorical variables. We also carried out additional analyses using CRP as a continuous variable (natural log-transformed).

### Outcomes

Our primary outcomes were depressive and anxiety symptoms occurring in the last 2 weeks as measured using the Patient Health Questionnaire (PHQ)-9 and the Generalised Anxiety Disorder (GAD)-7 questionnaire, respectively. Symptoms were coded as 0-3 depending on frequency. We created sum-scores for each scale, which were used as primary outcomes. Categorical diagnoses of probable depression and generalised anxiety disorder (GAD) were used as secondary outcomes, which were defined using commonly used cut-off criteria of PHQ-9≥10 and GAD-7≥10. See details in the Supplementary Appendix.

### Covariates

As covariates, we included age, sex, body mass index (BMI), smoking, alcohol use, physical activity, ethnicity, Townsend Deprivation Index (TDI), and diabetes and cardiovascular disease; see Supplementary Appendix for details.

### Statistical Analyses

Analyses were performed using Stata/SE 16.0 (Stata, College Station, TX). Baseline characteristics of participants were examined across CRP quintiles.

#### Association of CRP with depression and anxiety, linearity and sex difference

Linear regression was used to estimate the associations between CRP concentrations (quintiles or deciles) and depressive and anxiety symptom scores. For the purpose of interpretation, coefficient estimates were anti-log transformed to odds ratio and 95% confidence interval (CI). We adjusted regression models for age, sex, BMI, smoking, alcohol use, physical activity, ethnicity, TDI, and diabetes and cardiovascular disease.

To investigate the nature of associations with depressive and anxiety symptoms and any dose-response effect in greater detail, CRP concentrations were divided into deciles with deciles 2-10 compared with the lowest decile group (decile 1). Floating absolute risks were estimated, which were then plotted against the median CRP concentrations in each decile. We computed ORs for trend by using quintile number as predictor. We assessed potential quadratic associations by including a quadratic term (CRP-squared). We performed sex-stratified analyses and also tested for interaction between sex and CRP by including interaction terms in regression models. Lastly, we evaluated the influence of selection/collider bias for participation in the optional mental health survey using inverse probability weighted regression of the fully adjusted regression models of depression and anxiety outcomes on CRP; see Supplementary Methods for details.

#### Test for specificity vs commonality of association of CRP between depression and anxiety

We used bivariate probit regression to test for specificity of association of CRP between depression and anxiety using both continuous and categorical outcomes. Probit regression jointly modelled the outcomes of depression and anxiety with CRP, and then tested for equality of regression parameters expressing the effect of CRP on each outcome using the likelihood ratio test. We compared a model that allowed estimates to differ between outcomes with a model where estimates were constrained to be equal for both outcomes. Probit estimates were converted into ORs by multiplying probit parameters by 1.6.^19^ In addition, we adjusted the regression models of depression for anxiety (along with other covariates) and *vice versa* as additional tests for disorder specificity.

### Mendelian randomisation approach

#### Genotyping

We used genotyping data of 342,081 unrelated individuals of White ancestry; see Supplementary Methods for details on genotyping array, central and post-imputation quality control. We used a summary-based approach for MR analyses,^20^ so sample sizes differed for estimation of SNP-exposure and SNP-outcome associations. For estimation of SNP-outcome associations, sample sizes varied between 100,739-110,173 per outcome; see Supplementary Table 1 for sample sizes for SNP-exposure associations.

#### SNP selection

We selected genetic variants in the *CRP* and *IL-6 receptor* (*IL-6R*) gene regions previously shown to be associated with CRP or IL-6 concentrations (Supplementary Table 1).^21–24^ Genetic instruments differ in strength based on the precision with which they have been estimated in original GWAS studies. As instrument strength informs statistical power for MR analysis, we use genetic instruments from Georgakis *et al*.^21^ for primary MR analysis, which have the largest strength (Supplementary Table 1), and report results from other instruments^22–24^ as sensitivity analysis.

We extracted SNP-exposure estimates from previous reports to perform 2-sample MR analysis. Based on availability of CRP concentrations in the UK Biobank study, which can be used as downstream readout of IL-6 activity,^20^ we also estimated SNP-exposure associations (for 1-sample MR) and SNP-outcome associations, in the full sample and separately for men and women for sex-stratified MR; see details in Supplementary Methods.

#### Mendelian randomisation analyses

We performed MR analysis using inverse-variance weighted (IVW) regression of the genetic associations with the outcome on the genetic associations with the exposure.^20^ To evaluate the potential impact of selection/collider bias for participation in the optional mental health survey, we repeated IVW MR analyses with SNP-outcome associations obtained using inverse probability weighted regression.^25^ We also evaluated potential horizontal pleiotropy using Cochran’s *Q*.^20^ See details in Supplementary Appendix.

## RESULTS

### Baseline Characteristics

In 146,954 participants (43.6% men), mean age at recruitment was 56.5 (SD=7.8) years. Median CRP concentration was 1.15 mg/L (IQR=0.58-2.38 mg/L). Table 1 shows characteristics of study participants by CRP quintiles.

**Table 1.** Baseline characteristics of study participants by quintiles of CRP levels in the UK Biobank cohort (n=146,954)

| Study characteristics | Q1<br>(n=34,787) | Q2<br>(n=32,125) | Q3<br>(n=29,113) | Q4<br>(n=26,733) | Q5<br>(n=24,196) | P value |
| --- | --- | --- | --- | --- | --- | --- |
| CRP (mg/L) median (range) | 0.36 (0.08-0.55) | 0.77 (0.56-1.02) | 1.33 (1.03-1.75) | 2.33 (1.76-3.33) | 5.42 (3.34-78.22) | <0.001 |
| Age (years) | 54.3 (7.8) | 55.82 (7.7) | 56.5 (7.6) | 56.9 (7.6) | 56.6 (7.7) | <0.001 |
| Women (%) | 20262 (58.3) | 17255 (53.7) | 15588 (53.5) | 14867 (55.6) | 14931 (61.7) | <0.001 |
| White ethnicity (%) | 33601 (96.6) | 31166 (97.0) | 28228 (97.0) | 25907 (96.9) | 23399 (96.7) | <0.001 |
| TDI, median (SD) | -1.7 (2.8) | -1.8 (2.8) | -1.8 (2.8) | -1.7 (2.8) | -1.5 (2.9) | <0.001 |
| BMI (kg/m<sup>2</sup>) | 24.1 (3.1) | 25.8 (3.4) | 27.0 (3.9) | 28.2 (4.3) | 30.1 (5.8) | <0.001 |
| Smoking status (%) |  |  |  |  |  |  |
| Never | 21603 (62.1) | 18927 (58.9) | 16509 (56.7) | 14722 (55.1) | 12555 (51.9) |  |
| Current | 1965 (5.7) | 1981 (6.2) | 2057 (7.1) | 2162 (8.1) | 2418 (10.0) |  |
| Ex-smokers | 11157 (32.1) | 11138 (34.7) | 10484 (36.0) | 9783 (36.6) | 9163 (37.9) | <0.001 |
| Alcohol status (%) |  |  |  |  |  |  |
| Never/Ex | 1743 (5.0) | 1581 (4.9) | 1578 (5.4) | 1633 (6.1) | 1659 (6.9) |  |
| Occasional (≤ 3 times per week) | 14376 (41.3) | 13856 (43.2) | 13052 (44.8) | 12719 (47.6) | 12184 (50.4) |  |
| Regular (> 3 times per week) | 18657 (53.7) | 16677 (51.9) | 14475 (49.7) | 12369 (46.3) | 10342 (42.8) | <0.001 |
| Physical activity (%) |  |  |  |  |  |  |
| Inactivity | 27490 (90.0) | 24961 (80.1) | 22180 (79.1) | 19756 (77.7) | 16816 (74.9) |  |
| Moderately inactive | 1350 (4.0) | 1548 (5.0) | 1633 (5.8) | 1742 (6.9) | 1969 (8.8) |  |
| Moderately active | 4342 (12.8) | 3881 (12.5) | 3443 (12.3) | 3206 (12.6) | 2967 (13.2) |  |
| Active | 779 (2.3) | 778 (2.5) | 780 (2.8) | 722 (2.8) | 711 (3.2) | <0.001 |
| Diabetes (%) | 780 (2.2) | 881 (2.7) | 983 (3.4) | 1022 (3.8) | 1210 (5.0) | <0.001 |
| Cardiovascular disease (%) | 1029 (3.0) | 1093 (3.4) | 1076 (3.7) | 1035 (3.9) | 973 (4.0) | <0.001 |

*Note:* Differences were estimated using mean and SD for continuous variables, with p-values from ANOVA test, or using number and percent for categorical variables, with $\chi^2$ test.

### Association of CRP Concentration with Depressive and Anxiety Symptom Scores

Results for associations of CRP with depressive and anxiety symptoms are presented in Figure 1 across different CRP deciles in the total sample, and for women and men separately in Supplementary Figures 1 and 2. CRP was associated with depressive and anxiety symptoms (Supplementary Tables 2 & 3)

**Figure 1.**
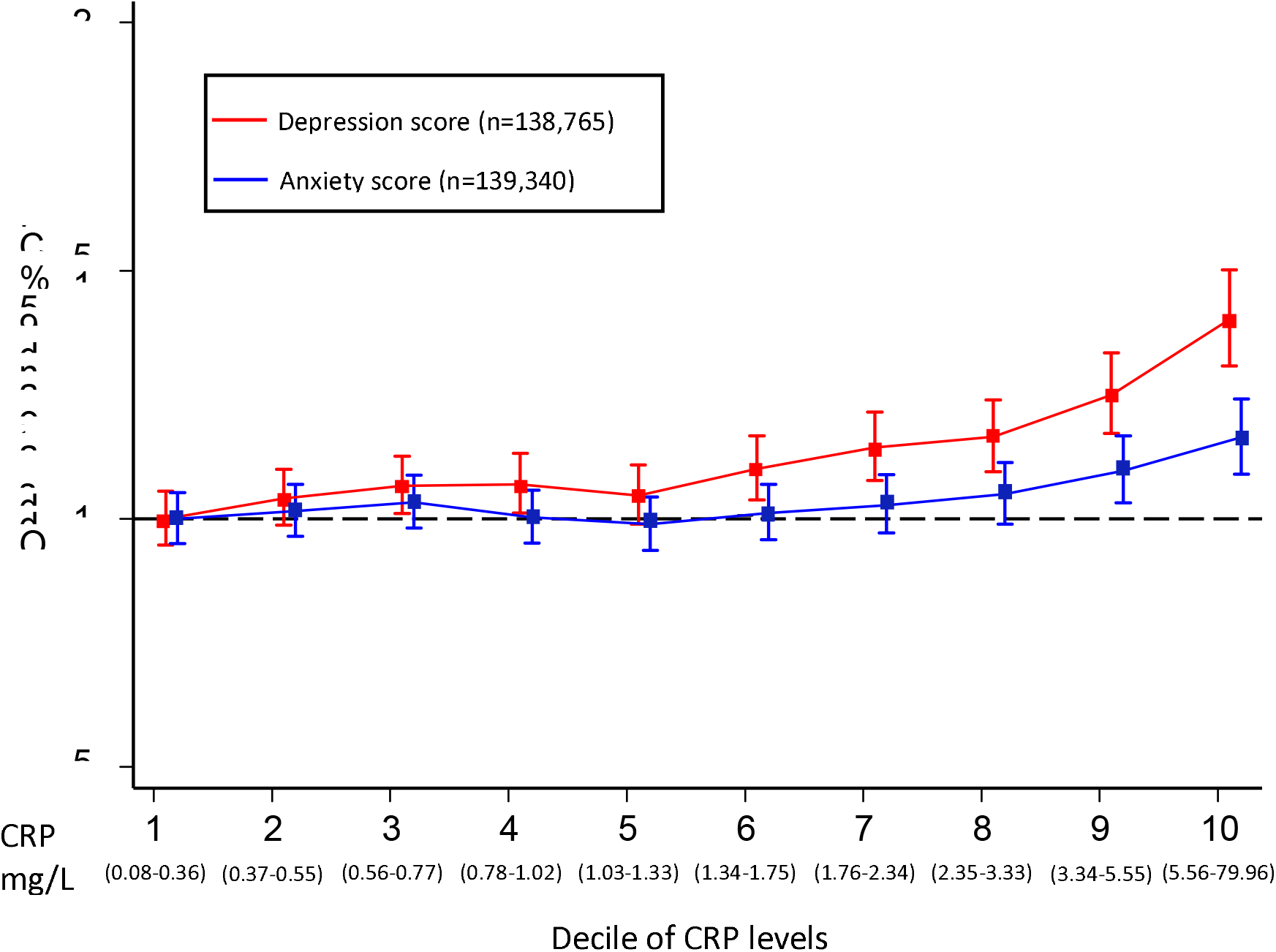
Odds ratios for higher depressive and anxiety symptom scores per decile of CRP levels in the UK Biobank cohort. CRP: C-reactive protein; Confidence intervals (CIs) were calculated using a floating absolute risk technique; Odds ratios were adjusted for age, sex, BMI, smoking status, alcohol intake, physical activity, TDI, ethnic group, diabetes and cardiovascular disease; red: depression score; blue: anxiety score

Using CRP as a continuous variable, the adjusted OR for higher depressive symptom score per-unit increase in CRP was 1.09 (95% CI, 1.06-1.11). Using CRP as a categorical variable, the adjusted OR for higher depressive symptom score for participants in the top, compared with bottom, quintile of CRP was 1.29 (95% CI, 1.21-1.38). Inverse probability weighted regression analyses of depressive symptoms did not suggest that results were affected by collider bias, as the adjusted OR=1.31 (95% CI, 1.22-1.41) for participants in the top, compared with bottom, quintile of CRP was similar.

Using CRP as a continuous variable, the adjusted OR for higher anxiety symptom score per-unit increase in CRP was 1.03 (95% CI, 1.02-1.05). Using CRP as a categorical variable, the adjusted OR for higher anxiety symptom score for participants in the top, compared with bottom, quintile of CRP was 1.12 (95% CI, 1.05-1.19). Again, evidence did not suggest results were affected by collider bias with similar OR of 1.12 (95% CI, 1.05-1.20) in sensitivity analyses.

### Association of CRP Concentration with Probable Diagnoses of Depression and GAD

CRP was associated with probable diagnosis of depression (Table 2). Using CRP as a continuous variable, the adjusted OR for depression per-unit increase in CRP was 1.09 (95% CI, 1.06-1.11). Using CRP as a categorical variable, the adjusted OR for depression for participants in the top, compared with bottom, quintile of CRP was 1.29 (95% CI, 1.18-1.40). Evidence did not suggest results were affected by collider bias with similar OR of 1.29 (95% CI, 1.18-1.41) in sensitivity analyses.

**Table 2.** Association of C-reactive protein levels with probable diagnosis of depression in the UK Biobank cohort.

|  | log CRP as continuous variable | CRP Q1 (n=34,372) | CRP Q2 (n=31,704) | CRP Q3 (n=28,714) | CRP Q4 (n=26,350) | CRP Q5 (n=23,750) | Per-Q effect | P-value for trend |
| --- | --- | --- | --- | --- | --- | --- | --- | --- |
| **All participants (cases = 8,888; controls = 145,468)** |  |  |  |  |  |  |  |  |
| Model 1 (n=144890) | 1.27 (1.24-1.29) | 1 [reference] | 1.11 (1.03-1.19) | 1.19 (1.10-1.28) | 1.44 (1.34-1.54) | 2.05 (1.91-2.20) | 1.19 (1.17-1.21) | <0.001 |
| Model 2 (n=144600) | 1.12 (1.09-1.15) | 1 [reference] | 1.08 (1.00-1.16) | 1.10 (1.02-1.18) | 1.22 (1.13-1.31) | 1.41 (1.31-1.53) | 1.09 (1.07-1.10) | <0.001 |
| Model 3 (n=138766) | 1.09 (1.06-1.11) | 1 [reference] | 1.07 (0.99-1.15) | 1.08 (1.00-1.17) | 1.16 (1.07-1.26) | 1.28 (1.18-1.39) | 1.06 (1.04-1.08) | <0.001 |
| Model 4 (n=138765) | 1.09 (1.06-1.11) | 1 [reference] | 1.07 (0.99-1.16) | 1.08 (1.00-1.17) | 1.16 (1.07-1.26) | 1.29 (1.18-1.40) | 1.06 (1.04-1.08) | <0.001 |
| **Women (cases = 5,641; controls = 81,562)** |  |  |  |  |  |  |  |  |
| Model 1 (n=81610) | 1.28 (1.25-1.32) | 1 [reference] | 1.06 (0.96-1.16) | 1.22 (1.12-1.34) | 1.40 (1.28-1.53) | 2.11 (1.94-2.29) | 1.20 (1.18-1.23) | <0.001 |
| Model 2 (n=81454) | 1.12 (1.08-1.15) | 1 [reference] | 1.03 (0.94-1.13) | 1.13 (1.03-1.24) | 1.18 (1.07-1.30) | 1.41 (1.27-1.55) | 1.09 (1.06-1.11) | <0.001 |
| Model 3 (n=77818) | 1.10 (1.06-1.14) | 1 [reference] | 1.02 (0.93-1.13) | 1.13 (1.02-1.25) | 1.17 (1.06-1.30) | 1.33 (1.20-1.48) | 1.07 (1.05-1.10) | <0.001 |
| Model 4 (n=77818) | 1.10 (1.06-1.13) | 1 [reference] | 1.02 (0.93-1.13) | 1.13 (1.02-1.25) | 1.17 (1.06-1.30) | 1.33 (1.20-1.48) | 1.07 (1.05-1.10) | <0.001 |
| **Men (cases = 3,247; controls = 63,906)** |  |  |  |  |  |  |  |  |
| Model 1 (n=63280) | 1.22 (1.18-1.27) | 1 [reference] | 1.23 (1.09-1.38) | 1.17 (1.04-1.32) | 1.53 (1.36-1.72) | 1.87 (1.66-2.11) | 1.16 (1.13-1.19) | <0.001 |
| Model 2 (n=63146) | 1.13 (1.08-1.17) | 1 [reference] | 1.16 (1.03-1.30) | 1.04 (0.92-1.18) | 1.27 (1.12-1.43) | 1.44 (1.27-1.64) | 1.08 (1.05-1.12) | <0.001 |
| Model 3 (n=60948) | 1.07 (1.02-1.11) | 1 [reference] | 1.12 (0.99-1.27) | 1.00 (0.88-1.14) | 1.14 (1.00-1.29) | 1.21 (1.06-1.39) | 1.04 (1.01-1.07) | 0.02 |
| Model 4 (n=60947) | 1.07 (1.03-1.12) | 1 [reference] | 1.13 (1.00-1.28) | 1.01 (0.89-1.15) | 1.15 (1.01-1.31) | 1.23 (1.07-1.41) | 1.04 (1.01-1.07) | 0.01 |

Data show OR and 95% CIs unless otherwise indicated. P for trend is from regression models with quintiles. Model 1, unadjusted; model 2, adjusted for age, sex, and BMI (body mass index); model 3, model 2 additionally adjusted for smoking, alcohol, physical activity, ethnicity, and TDI (Townsend deprivation index at recruitment); model 4, model 3 additionally adjusted for diabetes and cardiovascular disease; \*: CRP concentration was log transformed; Median CRP level was 1.15 mg/L (range 0.08-78.22 mg/L)

CRP was associated with probable diagnosis of GAD (Table 3). Using CRP as a continuous variable, the adjusted OR for GAD per-unit increase in CRP was 1.05 (95% CI, 1.02-1.08). Using CRP as a categorical variable, the adjusted OR for GAD for participants in the top, compared with bottom, quintile of CRP was 1.15 (95% CI, 1.05-1.26). Again, evidence did not support collider bias as likely explanation with similar OR of 1.13 (95% CI, 1.02-1.24) in sensitivity analyses.

**Table 3.** Association of C-reactive protein levels with probable GAD diagnosis in the UK Biobank cohort.

|  | log CRP as continuous variable | CRP Q1 (n=34,499) | CRP Q2 (n=31,809) | CRP Q3 (n=28,829) | CRP Q4 (n=26,451) | CRP Q5 (n=23,950) | Per-Q effect | P for trend |
| --- | --- | --- | --- | --- | --- | --- | --- | --- |
| **All participants (cases = 6,395; controls = 139,143)** |  |  |  |  |  |  |  |  |
| Model 1 (n=145,538) | 1.11 (1.08-1.14) | 1 [reference] | 0.95 (0.88-1.03) | 0.95 (0.88-1.03) | 1.05 (0.97-1.13) | 1.38 (1.28-1.49) | 1.08 (1.06-1.10) | <0.001 |
| Model 2 (n=145,239) | 1.07 (1.04-1.10) | 1 [reference] | 0.99 (0.91-1.07) | 0.99 (0.91-1.07) | 1.05 (0.97-1.14) | 1.24 (1.14-1.36) | 1.05 (1.03-1.07) | <0.001 |
| Model 3 (n=139,341) | 1.05 (1.02-1.08) | 1 [reference] | 0.97 (0.90-1.06) | 0.99 (0.91-1.07) | 1.02 (0.94-1.12) | 1.15 (1.05-1.26) | 1.03 (1.01-1.05) | 0.004 |
| Model 4 (n=139,340) | 1.05 (1.02-1.08) | 1 [reference] | 0.98 (0.90-1.06) | 0.98 (0.90-1.07) | 1.02 (0.94-1.11) | 1.15 (1.05-1.26) | 1.03 (1.01-1.05) | 0.005 |
| **Women (cases = 4,247; controls = 77,717)** |  |  |  |  |  |  |  |  |
| Model 1 (n=81,964) | 1.10 (1.07-1.13) | 1 [reference] | 0.97 (0.88-1.07) | 0.95 (0.86-1.05) | 1.03 (0.93-1.13) | 1.38 (1.26-1.51) | 1.07 (1.05-1.10) | <0.001 |
| Model 2 (n=81,799) | 1.08 (1.04-1.11) | 1 [reference] | 1.00 (0.91-1.10) | 0.98 (0.88-1.08) | 1.05 (0.94-1.16) | 1.29 (1.16-1.43) | 1.05 (1.03-1.08) | <0.001 |
| Model 3 (n=78,110) | 1.07 (1.03-1.10) | 1 [reference] | 0.98 (0.89-1.09) | 0.99 (0.90-1.10) | 1.04 (0.94-1.16) | 1.23 (1.10-1.38) | 1.05 (1.02-1.07) | 0.001 |
| Model 4 (n=78,110) | 1.06 (1.03-1.10) | 1 [reference] | 0.99 (0.89-1.09) | 0.99 (0.89-1.10) | 1.04 (0.93-1.16) | 1.23 (1.10-1.37) | 1.05 (1.02-1.07) | 0.001 |
| **Men (cases = 2,148; controls = 61,426)** |  |  |  |  |  |  |  |  |
| Model 1 (n=63,574) | 1.10 (1.06-1.16) | 1 [reference] | 0.97 (0.85-1.11) | 1.02 (0.89-1.17) | 1.12 (0.98-1.28) | 1.33 (1.16-1.53) | 1.07 (1.04-1.11) | <0.001 |
| Model 2 (n=63,440) | 1.07 (1.02-1.12) | 1 [reference] | 0.97 (0.85-1.11) | 1.02 (0.89-1.17) | 1.12 (0.98-1.28) | 1.33 (1.16-1.53) | 1.04 (1.01-1.08) | 0.018 |
| Model 3 (n=61,231) | 1.02 (0.98-1.07) | 1 [reference] | 0.94 (0.82-1.08) | 0.95 (0.83-1.10) | 0.97 (0.84-1.13) | 1.02 (0.87-1.20) | 1.01 (0.97-1.04) | 0.74 |
| Model 4 (n=61,230) | 1.02 (0.98-1.07) | 1 [reference] | 0.95 (0.82-1.08) | 0.95 (0.83-1.10) | 0.97 (0.84-1.13) | 1.02 (0.87-1.20) | 1.01 (0.97-1.04) | 0.74 |

*Note:* Data show ORs and 95% CIs unless otherwise indicated. P for trend is from regression models with quintiles. Model 1, unadjusted; model 2, adjusted for age, sex, and BMI (body mass index); model 3, model 2 additionally adjusted for smoking, alcohol, physical activity, ethnicity, and TDI (Townsend deprivation index at recruitment); model 4, model 3 additionally adjusted for diabetes and cardiovascular disease; \*: CRP concentration was log transformed; Median CRP level was 1.33 mg/L (range 0.08-79.96 mg/L).

### Test for specificity vs commonality of association of CRP with depression and anxiety

In bi-variate probit regression analysis, we found evidence for a stronger association of CRP with depressive symptoms (OR=1.014; 95% CI, 1.011-1.017) than anxiety symptoms (OR=1.004; 95% CI, 1.002-1.007). Results for probit regression using probable diagnoses of depression and GAD as outcomes were similar (see Supplementary Results).

In regression analyses, evidence for association of CRP with depression symptoms remained after adjusting for anxiety symptoms (OR=1.06; 95% CI, 1.05-1.08), but the association of CRP with anxiety symptoms switched its valence after adjusting for depressive symptoms (OR=0.98; 95% CI, 0.97-0.99).

### Linearity of association

Evidence was compatible with linear associations of CRP with both depression and anxiety across all analyses using symptom scores and probable diagnoses as outcomes (*P*-value for all quadratic terms >0.05).

### Examination of potential sex difference

In sex-stratified analyses, point estimates were larger for women than men for both depression and anxiety symptom outcomes (Supplementary Tables 2-3, Supplementary Figures 1-2). However, evidence for an interaction between CRP and sex was present only for depressive symptoms (adjusted OR_women_=1.35; 95%CI, 1.23-1.48; adjusted OR_men_=1.21; 95%CI, 1.10-1.33; *P*-value for interaction term=0.032). For categorical outcomes, point estimates were larger for women for probable GAD (Tables 2-3), but evidence did not support interaction for either outcomes (all *P*>0.2).

### Results for Mendelian randomization analyses

Genetically-predicted concentration/activity of IL-6 and CRP were associated with both depression and anxiety. However, these associations differed with regards to direction of association (i.e., increased vs decreased risk), particular outcome definition, and sex. Table 4 shows results for IVW MR analyses based on Georgakis *et al*.^21^ genetic instruments for CRP and IL-6.

**Table 4.** IVW Mendelian randomisation analysis of association of IL-6 and CRP with depression and anxiety.

|  | Depression Symptom Score |  | Probable depression |  | Anxiety Symptom Score |  | Probable GAD |  |
| --- | --- | --- | --- | --- | --- | --- | --- | --- |
| Model | OR (95% CI) | P-value | OR (95% CI) | P-value | OR (95% CI) | P-value | OR (95% CI) | P-value |
| **CRP** |  |  |  |  |  |  |  |  |
| 2-Sample MR | 0.88 (0.80-0.98) | 0.020 | 0.95 (0.85-1.07) | 0.424 | 0.87 (0.80-0.95) | 0.003 | 0.82 (0.72-0.94) | 0.004 |
| 1-Sample MR | 0.89 (0.79-1.00) | 0.055 | 1.01 (0.88-1.14) | 0.939 | 0.88 (0.79-0.97) | 0.008 | 0.84 (0.73-0.98) | 0.027 |
| Women | 0.98 (0.85-1.12) | 0.754 | 1.12 (0.96-1.30) | 0.152 | 0.86 (0.76-0.98) | 0.023 | 0.85 (0.72-1.01) | 0.059 |
| Men | 0.78 (0.63-0.96) | 0.018 | 0.84 (0.66-1.06) | 0.138 | 0.91 (0.78-1.05) | 0.192 | 0.83 (0.62-1.11) | 0.209 |
| **IL-6** |  |  |  |  |  |  |  |  |
| 2-Sample MR | 1.34 (1.05-1.72) | 0.019 | 1.15 (0.86-1.54) | 0.340 | 1.13 (0.91-1.41) | 0.269 | 1.24 (0.89-1.73) | 0.194 |
| 1-Sample MR | 1.32 (1.03-1.67) | 0.025 | 1.18 (0.89-1.56) | 0.246 | 1.11 (0.90-1.37) | 0.313 | 1.18 (0.86-1.62) | 0.297 |
| Women | 1.42 (1.01-1.97) | 0.041 | 1.46 (1.00-2.13) | 0.048 | 1.15 (0.85-1.56) | 0.362 | 1.51 (1.01-2.25) | 0.044 |
| Men | 1.24 (0.88-1.74) | 0.218 | 0.86 (0.54-1.37) | 0.516 | 1.08 (0.79-1.47) | 0.636 | 0.79 (0.47-1.33) | 0.385 |

*Note:* Estimates for men and women are based on sex-stratified 1-sample MR analyses.

For CRP, per-unit increase in genetically-predicted concentrations of log-transformed CRP was associated with lower risk for depressive symptoms (1-sample MR: OR=0.89; 95% CI, 0.79-1.00; 2-sample MR: OR=0.88; 95% CI, 0.80-0.98), and lower risk for anxiety symptoms (1-sample MR: OR=0.88; 95% CI, 0.79-0.97; 2-sample MR: OR=0.87; 95% CI, 0.80-0.95).

Using the categorical outcomes, MR analyses also showed that increased genetically-predicted CRP was associated with lower risk for probable GAD, but point estimates for probable depression were close to one (Table 4). In sex-stratified MR analyses, higher genetically predicted CRP concentrations were associated with relatively lower risk for depressive symptoms in men, and with relatively lower risk for anxiety symptoms in women.

For IL-6, per-unit increase in higher genetically-predicted IL-6 activity was associated with increased risk for depressive symptoms (1-sample MR: OR=1.32, 95% CI 1.03-1.67; 2-sample MR: OR=1.34, 95% CI 1.05-1.72), but not with probable depression or either anxiety outcome. In sex-stratified MR analyses, we found evidence that higher genetically-predicted IL-6 activity was associated with increased risk for depressive symptoms, probable depression, and probable GAD in women only.

MR analyses using alternative genetic instruments were directionally consistent with these results, albeit with larger confidence intervals possibly due to the lower statistical power for these instruments (Supplementary Table 4). Results for sensitivity analyses evaluating the impact of selection/collider bias were similar to main IVW analyses (Supplementary Table 5).

Evidence did not suggest directional horizontal pleiotropy was a likely explanation for any of the IVW MR results as assessed using Cochran’s *Q* (Supplementary Table 6).

## DISCUSSION

Based on data from the UK Biobank cohort, a large general population cohort, we report that circulating CRP concentrations are associated with depressive and anxiety symptoms and with probable diagnoses of depression and GAD in a linear, dose-response fashion. At the same time, we show evidence for disorder-specificity suggesting that CRP is more strongly associated with depression compared to anxiety. We also found some evidence for sex-specificity. CRP was more strongly associated with depression in women than in men. Using MR analyses, we provide evidence that higher IL-6 activity could represent a potential causal factor increasing depression, while higher CRP concentrations could potentially be protective for depression and anxiety.

### Associations of inflammation with depression and anxiety

Although inflammation was associated with both depression and anxiety, we report stronger associations for depression outcomes indicating disorder-specificity. This aligns with meta-analyses of case-control studies showing higher concentrations of CRP and other inflammatory markers in depression,^4^ while there are relatively fewer studies suggesting this for anxiety.^26^ Cohort studies of affective symptoms also suggest that circulating IL-6 and CRP concentrations are predominantly associated with depressive rather than anxiety symptoms.^27^ Together, current evidence is consistent with the idea that systemic inflammation may be particularly relevant for depression rather than anxiety disorders.

Our results also indicate some evidence for sex-specificity. Associations of CRP concentrations with depression and anxiety were mostly stronger in women than men. Results for sex-stratified MR analyses also suggest that IL-6 could be a risk factor for depression specifically for women. However, existing evidence on potential sex-difference for associations between inflammatory makers and depression are mixed. A previous meta-analysis reported no sex-specificity of the association between CRP and depression.^3^ In contrast, two recent studies reported that IL-6 was associated with depressive symptom chronicity and treatment response specifically in women.^28,29^ Atypical depression, which is characterised by immuno-metabolic dysregulation, has also been reported to be more common in women.^30^ Hitherto most studies have considered sex as a covariate. Further research is needed to replicate our findings regarding potential sex-specificity.

Our findings support RCTs of immunotherapies targeting the IL-6/IL-6R pathway for depression. Anti-inflammatory treatments have been shown to exhibit antidepressant activity in chronic inflammatory illnesses.^31–33^ In depression, initial results suggest that these drugs may be useful for patients with evidence of inflammation and inflammation-related risk factors.^34,35^ This hypothesis is now being investigated in ongoing RCTs that are selecting patients based on evidence of inflammation and inflammation-related phenotypes.^6^ The present study further highlights characteristics associated with inflammation, e.g., female sex, to inform stratified patient selection in future clinical trials.

### Potential interpretations for divergent effects of CRP and IL-6

Using genetic variants in the *IL-6R* and *CRP* gene loci, we have found that higher IL-6 activity was associated with increased risk of depression, but higher CRP levels were associated with decreased risk of depression. These findings are intriguing because IL-6 signalling is a key driver of CRP response,^36^ and so we would expect both to affect depression risk in a comparable way. One potential explanation could be that IL-6 classic and trans-signalling have divergent effects on depression risk. We have illustrated this hypothesis in Supplementary Figure 3, which describes IL-6 signalling pathways and a Directed Acyclic Graph of these pathways incorporating our MR results.

In brief, IL-6 classic signalling occurs via its action on membrane-bound IL-6 receptors (IL-6Rs) expressed by limited cell types. IL-6 also binds with circulating soluble IL-6R (sIL-6R) to form an IL-6-sIL-6R complex, which then activates IL-6 signalling by binding with the ubiquitous glycoprotein 130 on other cells that naturally lack IL-6Rs. This is called IL-6 trans-signalling, which is thought to underlie pro-inflammatory pathological effects of IL-6 in chronic inflammatory diseases.^36^

Mechanistically, observed increased depression risk conferred by *IL6R* SNPs that increase CRP levels^21^ could happen as a result of either increased IL-6 classic or trans-signalling. Our results indicate that it may be due to increased trans-signalling, because we also see that *CRP* SNPs that increase CRP levels^21^ are protective for depression. It is well-known that CRP is mainly produced by hepatocytes as a result of increased IL-6 classic signalling.^36^

While the MR approach can provide evidence supporting causality, as we do here for IL-6 and depression, disentangling the issue of IL-6 classic vs trans-signalling is beyond the scope of population genomics approaches as full effects of genetic variants used are unknown. The field now requires experimental studies of IL-6 modulation in humans and animals to further examine causality, pathogenic mechanisms, and therapeutic potential of anti-IL-6 and other immunotherapies for depression. Findings from these studies may help to devise more targeted IL-6 pathway-specific interventions.

### Strengths and Limitations

Strengths of the work include use of a large population-based sample, a range of affective symptoms, complementary analysis using protein levels and genetic variants. We assessed reproducibility and strength of association using different outcomes and sex-stratified analysis, evidence of linearity and potential causality of associations. Limitations of the work include focus on symptom score/probable diagnosis. Depression is a phenotypically heterogeneous syndrome and previous studies have reported that inflammation may be associated with specific symptoms, such as fatigue, changes in appetite and sleep, and suicidality.^16,27,30^ Although there was little evidence that associations of CRP with depression and anxiety could be due to selection/collider bias into the optional UK Biobank Mental Health Survey, selection/collider bias for participation in the UK Biobank cohort itself would likely be larger and remains a possible explanation for our findings that we could not explore. Third, MR findings were based on a subgroup of individuals of European ancestry, which is a common issue in genetic studies, warranting replication in other ethnic groups. Finally, IL-6 was not measured in the UK Biobank cohort, so we were unable to assess associations of serum IL-6 concentrations with depression and anxiety.

## Conclusions

We report evidence for associations of higher CRP concentrations with depression and anxiety, which are stronger for depression than for anxiety and, although less consistently, for women than for men. Findings from MR analyses are consistent with a causal role of altered activity of the IL-6/IL-6R pathway in depression, suggesting that this pathway could be a promising, new therapeutic target for depression. Human and animal experimental studies are required to elucidate mechanisms for divergent effects for CRP and IL-6 on illness risk, as this may help to devise more targeted interventions.

## Supporting information

STROBE checklist

## Data Availability

UK Biobank data can be accessed through formal application to the cohort. GWAS summary data used as part of this report are freely available online or can be requested for CRP from the CHARGE inflammation working group. Genetic instrument estimates and scripts for MR processing and analysis are made available online for full reproducibility under https://osf.io/apme9/.

## ACKNOWLEDGMENTS

This work was supported by a Data Science Award from the MQ: Transforming Mental Health (grant code: MQDS17/40) to GMK and PBJ, which also supported ZY. GMK also acknowledges funding support from the Wellcome Trust (grant code: 201486/Z/16/Z), the Medical Research Council UK (grant code: MC_PC_17213 and MR/S037675/1), and the BMA Foundation (J Moulton grant 2019). NK and SM are supported by the International Max Planck Research School of Translational Psychiatry (IMPRS-TP). GDS works in the Medical Research Council Integrative Epidemiology Unit at the University of Bristol, which is supported by the Medical Research Council (MC_UU_00011/1).

