## Supplementary Material for "Role of Inflammation in Depression and Anxiety: Tests for Disorder Specificity, Linearity and Potential Causality of Association in the UK Biobank"

### SUPPLEMENTARY METHODS

#### Study Population

The UK Biobank study was approved by the UK Biobank’s research ethics committee and Human Tissue Authority research tissue bank. An independent Ethics and Governance Council oversees adherence to the Ethics and Governance Framework and provides advice on the interests of research participants and the general public in relation to UK Biobank.^1^ This project was approved by the UK Biobank study (ref no. 26999). Informed consent was obtained from all participants.

#### Outcomes

##### Depressive Symptom Score

Depressive symptoms occurring in the last two weeks were measured using the patient health questionnaire (PHQ-9), a validated tool for assessing depressive symptoms,^2^ which was administered online, as part of a follow-up mental health survey completed by up to 157,115 participants.^3,4^ The PHQ-9 comprises nine depressive symptoms, including feelings of inadequacy, concentration problems, low mood, changes in appetite, thoughts of suicide/self-harm, anhedonia, sleep problems, psychomotor agitation/retardation, and fatigue. Each item was coded as not at all (0), several days (1), more than half the days (2), nearly every day (3), or prefer not to answer, which was excluded from analysis. A total depressive symptom score was created by summing responses to individual items resulting in a total score of 0-27.

##### Anxiety Symptom Score

Anxiety symptoms were assessed using the validated Generalised Anxiety Disorder questionnaire (GAD-7),^5^ also administered as part of the follow-up mental health survey. It assesses seven symptoms including irritability, feelings of nervousness/anxiety, inability to stop or control worrying, feelings of foreboding, trouble relaxing, restlessness, and worrying too much about different things. Items were coded in the same way as depressive symptoms (above). A total score was created by summing individual items resulting in a total score of 0-21.

##### Probable Diagnosis of Depression

Probable diagnosis of depression was defined based on a total PHQ-9 score of ≥10. This cut-off score has been reported to have 85% sensitivity and 89% specificity for the diagnosis of depression. Those with scores ≤9 were used as comparison group.^6^

##### Probable Diagnosis of Generalised Anxiety Disorder

Probable diagnosis of GAD was defined based on a total GAD-7 score of ≥10 that has been reported to identify cases of GAD with 89% sensitivity and 82% specificity.^5^ Those with scores ≤9 were used as comparison group.

#### Covariates

The covariates included age, sex, body mass index (BMI), smoking, alcohol use, physical activity, ethnicity, Townsend Deprivation Index (TDI), and diabetes and cardiovascular disease. Smoking status was categorized into four groups (current, previous, never, prefer not to answer); alcohol drinking categorized into three groups (regular (> 3 times/week), occasional (< 3 times/week), never); physical activity categorized into four groups (active, moderately active, moderately inactive, and inactive). Presence of diabetes or cardiovascular disease was self-reported. We dichotomised ethnicity as “White” versus “other ethnicity”. TDI is a continuous measure of material deprivation in a given area that incorporates information on employment status, car and home ownership, and household crowding.^7^ Higher scores represent greater deprivation.

#### Evaluation of Selection Bias onto the Association of CRP with depression and anxiety

Only approximately a third of UK Biobank participants took part in the optional follow-up mental health survey.^4^ We evaluated the influence of potential collider/selection bias using inverse probability weighted regression of depression and anxiety outcome variables on CRP.^8^ This approach essentially upweights participants who were less likely to participate in the follow-up mental health survey and downweighs participants who were more likely to participate. In detail, this sensitivity analysis followed three steps. First, we regressed mental health questionnaire participation (coded as 1=participated, 0=did not participate) on age, sex, BMI, and TDI as well as all possible variable interactions and quadratic and cubic effects for continuous variables. Second, we created inverse probability weights from this regression by dividing the mean proportion of mental health survey participation by the fitted values from this regression; this approach also stabilises the weights as described by Sayon-Orea *and colleagues*.^9^ Third, we repeated regression analyses of depression and anxiety outcome variables on CRP quintiles and adjusted for all covariates, but using the inverse probability weights as regression weightings.

#### Mendelian randomisation approach

##### Genotyping

Genotyping data are available for 502,520 participants in the UK Biobank study. Genotyping was performed using the Affymetrix UK BiLEVE Axiom array on an initial 500,00 participants; the remaining 450,000 participants were genotyped using the Affymetrix UK Biobank Axiom® array, which genotyped ~850,000 variants. The two arrays are similar (with >95% common content). For further details on genotyping and quality control, see <https://www.ukbiobank.ac.uk/scientists-3/genetic-data/> and Bycroft *and colleagues*^10^.

In post-imputation quality control, the sample was further restricted to 342,081 individuals in control by removing individuals with non-British ancestry and individuals related up to third degree. SNP-based quality control criteria were minor allele frequency (MAF) <1%, imputation score <60%, genotyping missingness >2% and Hardy-Weinberg Equilibrium (HWE) test p-value <0.000001, which were applied using *QCTOOL* (<https://enkre.net/cgi-bin/code/qctool/dir?ci=trunk>).

##### SNP selection

For CRP, we defined genetic proxy instruments using 24 and 3 SNPs in the *CRP* gene region that exhibited genome-wide significant associations with CRP concentrations in two prior reports.^11,12^ For IL-6, we defined genetic proxy instruments using SNPs in the *IL-6R* gene. The first proxy instrument was based on 7 SNPs that exhibited genome-wide significant associations with CRP concentrations as a downstream readout of IL-6 activity.^12^ The second proxy instrument was based on 3 SNPs that showed genome-wide significant associations with IL-6 concentrations.^13^ Lastly, we also selected rs2228145 as a single-SNP instrument based on a prior report,^14^ which has been functionally implicated in IL-6 signalling and exhibited associations to psychiatric phenotypes in prior research.^15,16^

We extracted SNP-exposure estimates from previous reports to perform 2-sample MR analysis. Based on availability of CRP concentrations in UK Biobank, which can also be used to index IL-6 activity,^12^ we also estimated SNP-exposure estimates to perform 1-sample MR. These SNP-exposure estimates were obtained by regressing log(CRP) concentrations on individual SNPs while controlling for 20 principal genotype components, age, age^2^, sex, and age*sex interaction. SNP-outcome estimates were obtained with a matched linear/logistic regression approach for continuous and categorical depression and anxiety phenotypes. To perform sex-stratified analyses, we also estimated SNP-exposure and SNP-outcome associations separately for men and women, but removed sex-related covariates from regression models.

Supplementary Table 1 displays sample sizes and power of all included genetic instruments.

##### Supplementary Table 1. Genetic instruments and sample sizes used to estimate SNP-exposure and SNP-outcome associations

|  | **2-Sample MR** | | | | **1-Sample MR** | | | |
| --- | --- | --- | --- | --- | --- | --- | --- | --- |
| **Instrument** | **Phenotype** | **#SNPs** | **Instrument F-statistics^a^** | **Sample N^b^** | **Phenotype** | **#SNPs** | **Instrument F-statistics^a^** | **Sample N^b^** |
| ***CRP*** |  |  |  |  |  |  |  |  |
| Georgakis *et al.*^12^ | CRP | 24 | Min=55; Median=274; Max=3413 | 204.402 | CRP | 24 | Min=24; Median=160; Max=4573 | 304,610-325,441 |
| CCGC^11^ | CRP | 3 | Min=236; Median=649; Max=1349 | 38,573-105,476 | CRP | 4 | Min=1750; Median=2510; Max=4573 | 322,826-325,441 |
| ***IL-6*** |  |  |  |  |  |  |  |  |
| Georgakis *et al.*^12^ | CRP | 7 | Min=80; Median=138; Max=764 | 204.402 | CRP | 7 | Min=127; Median=175; Max=1509 | 299,591-325,441 |
| Swerdlow *et al.*^13^ | IL-6 | 3 | Min=8;  Median=14;  Max=16 | 4,462-4,479 | CRP | 3 | Min=618; Median=620; Max=1489 | 323,494-325,369 |
| Sarwar *et al.*^14^ | IL-6 | 1 | 397^c^ | 27.185 | CRP | 1 | 1510 | 325,441 |

*Note*: ^a^Instrument strength F-statistics are based on the formulae $R^{2}=2*MAF*\left( 1-MAF \right)*{beta}^{2}$, where MAF=Minor allele frequency, and $F= \frac{R^{2}* ( N-2 )}{1-R^{2}}$ ; as described in Shim *et al.*^17^ and Palmer *et al.*^18^ previously. ^b^Sample sizes are provided as range as this can be varying per SNP. If single values are presented, there is no variability. ^c^Since MAF/ effect allele frequency were not available, the approximate F-statistic was calculated as $F= \frac{{beta}^{2}}{{se}^{2}}$ as described in Rosa *et al.*^19^ and Pierce *et al.*^20^ previously.

##### Mendelian randomisation analyses

MR analyses were conducted using *R* software version 4.0.3.^21^ and the *TwoSampleMR* package.^22^

We performed MR analysis using inverse-variance weighted (IVW) regression of the genetic associations with the outcome on the genetic associations with the predictor.^23^ The MR estimates represent the odds ratio for the outcome per unit increase in inflammatory marker concentrations.

As depression and anxiety outcome variables were based on an optional follow-up mental health survey within the UK Biobank study,^4^ we performed additional MR analyses to evaluate the impact of selection/collider bias on MR analyses.^8^ To this end, we followed the same approach as for phenotypic analyses. We first performed a regression analysis of mental health survey participation on sex, age, BMI, and TDI as predictor variables including all possible predictor interactions and quadratic and cubic effects for age, sex, BMI, and TDI (n=498,781). We then extracted stabilised inverse probabilities of participation,^9^ which were then used as weights in regression analyses to obtain SNP-outcome estimates for MR analyses. Of note, while this approach evaluates the impact of selection/collider bias posed by mental health survey participation, the impact of selection/collider bias through UK Biobank participation in general remains unknown.

Lastly, we evaluated the presence of potential horizontal pleiotropy using Cochran’s *Q*.^24^

### SUPPLEMENTARY RESULTS

#### Association of CRP Concentration with Depressive and Anxiety Symptom Scores

##### Supplementary Figure 1. Odds ratios for depression and anxiety scores per deciles of CRP levels in women in the UK Biobank cohort

.

5

1

1

.

5

2

O

d

d

s

r

a

t

i

o

(

9

5

%

C

I

)

Anxiety score

Depression score

1

2

3

4

5

6

7

8

9

10

Decile of CRP levels

(0.08-0.36)

(0.37-0.55)

(0.56-0.77)

(0.78-1.02)

(1.03-1.33)

(1.34-1.75)

(1.76-2.34)

(2.35-3.33)

(3.34-5.55)

(5.56-79.96)

CRP mg/L

CRP: C-reactive protein; Confidence intervals (CIs) were calculated using a floating absolute risk technique; Odds ratios were adjusted for age, sex, BMI, smoking status, alcohol intake, physical activity, TDI, ethnic group, diabetes and cardiovascular disease; black: depression and anxiety score; red: depression score; blue: anxiety score

##### Supplementary Figure 2. Odds ratios for depression and anxiety scores per deciles of CRP levels in men in the UK Biobank cohort

.

5

1

1

.

5

2

O

d

d

s

r

a

t

i

o

(

9

5

%

C

I

)

1

2

3

4

5

6

7

8

9

10

Decile of CRP levels

(0.08-0.36)

(0.37-0.55)

(0.56-0.77)

(0.78-1.02)

(1.03-1.33)

(1.34-1.75)

(1.76-2.34)

(2.35-3.33)

(3.34-5.55)

(5.56-79.96)

CRP mg/L

Anxiety score

Depression score

CRP: C-reactive protein; Confidence intervals (CIs) were calculated using a floating absolute risk technique; Odds ratios were adjusted for age, sex, BMI, smoking status, alcohol intake, physical activity, TDI, ethnic group, diabetes and cardiovascular disease; black: depression and anxiety score; red: depression score; blue: anxiety score

##### Supplementary Table 2. Association of C-reactive protein levels with depression score in the UK Biobank cohort

|  | **log CRP as continuous variable** | **Q1 (n=34,372)** | **Q2 (n=31,704)** | **Q3 (n=28,714)** | **Q4 (n=26,350)** | **Q5 (n=23,750)** | **Per-Q effect** | **P for trend** |
| --- | --- | --- | --- | --- | --- | --- | --- | --- |
| ***All participants*** |  |  |  |  |  |  |  |  |
| CRP (mg/L), median (range) | 0.14 (-2.53-4.36)* | 0.36 (0.08-0.55) | 0.77 (0.56-1.02) | 1.33 (1.03-1.75) | 2.33 (1.76-3.33) | 5.42 (3.34-78.22) |  |  |
| Model 1 (n=144,890) | 1.33 （1.30-1.35） | 1 [reference] | 1.10 (1.04-1.17) | 1.21 (1.14-1.28) | 1.53 (1.44-1.62) | 2.46 (2.32-2.62) | 1.23（1.21-1.24） | <0.001 |
| Model 2 (n=144,600) | 1.13 （1.11-1.15） | 1 [reference] | 1.05 (0.99-1.11) | 1.07 (1.01-1.13) | 1.19 (1.12-1.26) | 1.48 (1.38-1.58) | 1.09（1.07-1.11） | <0.001 |
| Model 3 (n=138,766) | 1.09 （1.06-1.11） | 1 [reference] | 1.04 (0.99-1.10) | 1.05 (0.99-1.11) | 1.12 (1.06-1.19) | 1.29 (1.21-1.37) | 1.06（1.04-1.07） | <0.001 |
| Model 4 (n=138,765) | 1.09 （1.06-1.11） | 1 [reference] | 1.05 (0.99-1.11) | 1.05 (0.99-1.11) | 1.13 (1.06-1.20) | 1.29 (1.21-1.38) | 1.06（1.04-1.07） | <0.001 |
| ***Women*** |  |  |  |  |  |  |  |  |
| Model 1 (n=81,610) | 1.38 （1.34-1.41） | 1 [reference] | 1.09 (1.01-1.18) | 1.25 (1.16-1.36) | 1.55 (1.43-1.67) | 2.77 (2.56-3.00) | 1.26 (1.24-1.28) | <0.001 |
| Model 2 (n=81,454) | 1.13 （1.10-1.16） | 1 [reference] | 1.02 (0.95-1.10) | 1.07 (0.99-1.16) | 1.15 (1.06-1.25) | 1.48 (1.36-1.63) | 1.09 (1.07-1.11) | <0.001 |
| Model 3 (n=77,818) | 1.10 （1.07-1.13） | 1 [reference] | 1.13 (1.04-1.23) | 1.08 (0.99-1.17) | 1.13 (1.04-1.23) | 1.36 (1.24-1.48) | 1.07 (1.05-1.09) | <0.001 |
| Model 4 (n=77,818) | 1.10 （1.07-1.13） | 1 [reference] | 1.03 (0.95-1.11) | 1.08 (0.99-1.17) | 1.13 (1.04-1.23) | 1.35 (1.23-1.48) | 1.07 (1.05-1.09) | <0.001 |
| ***Men*** |  |  |  |  |  |  |  |  |
| Model 1 (n=63,280) | 1.24 （1.21-1.27） | 1 [reference] | 1.18 (1.09-1.28) | 1.23 (1.13-1.33) | 1.55 (1.43-1.69) | 1.94 (1.77-2.13) | 1.17 (1.15-1.19) | <0.001 |
| Model 2 (n=63,146) | 1.13 （1.10-1.16） | 1 [reference] | 1.09 (1.01-1.18) | 1.06 (0.98-1.16) | 1.25 (1.14-1.36) | 1.45 (1.31-1.59) | 1.09 (1.06-1.11) | <0.001 |
| Model 3 (n=60,948) | 1.07 （1.04-1.10） | 1 [reference] | 1.07 (0.99-1.16) | 1.02 (0.94-1.11) | 1.13 (1.04-1.24) | 1.20 (1.10-1.32) | 1.04 (1.02-1.06) | <0.001 |
| Model 4 (n=60,947) | 1.07 （1.04-1.10） | 1 [reference] | 1.07 (0.99-1.16) | 1.03 (0.95-1.12) | 1.14 (1.05-1.25) | 1.21 (1.10-1.33) | 1.04 (1.02-1.07) | <0.001 |

*Note*: Data show ORs and 95% CIs unless otherwise indicated. P for trend is from regression models with quintiles. Model 1, unadjusted; model 2, adjusted for age, sex, and BMI (body mass index); model 3, model 2 additionally adjusted for smoking, alcohol, physical activity, ethnicity, and TDI (Townsend deprivation index at recruitment); model 4, model 3 additionally adjusted for diabetes and cardiovascular disease; *: CRP concentration was log transformed; Median CRP level was 1.15 mg/L (range 0.08-78.22 mg/L).

##### Supplementary Table 3. Association of C-reactive protein levels with anxiety score in the UK Biobank cohort

|  | **log CRP as continuous variable** | **Q1 (n=34,499)** | **Q2 (n=31,809)** | **Q3 (n=28,829)** | **Q4 (n=26,451)** | **Q5 (n=23,950)** | **Per-Q effect** | **P for trend** |
| --- | --- | --- | --- | --- | --- | --- | --- | --- |
| ***All participants*** |  |  |  |  |  |  |  |  |
| CRP (mg/L), median (range) | 0.14 (-2.53-4.36)* | 0.36 (0.08-0.55) | 0.77 (0.56-1.02) | 1.33 (1.03-1.75) | 2.33 (1.76-3.33) | 5.43 (3.34-78.22) |  |  |
| Model 1 (n=145,538) | 1.08 (1.06-1.09) | 1 [reference] | 0.95 (0.90-1.00) | 0.93 (0.88-0.98) | 1.02 (0.96-1.08) | 1.30 (1.23-1.38) | 1.06 (1.04-1.07) | <0.001 |
| Model 2 (n=145,239) | 1.06 (1.04-1.08) | 1 [reference] | 1.01 (0.96-1.07) | 0.99 (0.94-1.05) | 1.06 (1.00-1.12) | 1.21 (1.14-1.29) | 1.04 (1.03-1.05) | <0.001 |
| Model 3 (n=139,341) | 1.04 (1.02-1.06) | 1 [reference] | 1.01 (0.96-1.06) | 0.99 (0.94-1.05) | 1.03 (0.97-1.09) | 1.12 (1.05-1.19) | 1.02 (1.01-1.04) | 0.001 |
| Model 4 (n=139,340) | 1.03 (1.02-1.05) | 1 [reference] | 1.01 (0.96-1.06) | 0.99 (0.94-1.05) | 1.03 (0.97-1.09) | 1.12 (1.05-1.19) | 1.02 (1.01-1.04) | 0.002 |
| ***Women*** |  |  |  |  |  |  |  |  |
| Model 1 (n=81,964) | 1.07 (1.05-1.09) | 1 [reference] | 0.97 (0.90-1.04) | 0.95 (0.88-1.02) | 0.99 (0.91-1.06) | 1.30 (1.20-1.40) | 1.05 (1.03-1.07) | <0.001 |
| Model 2 (n=81,799) | 1.06 (1.03-1.09) | 1 [reference] | 1.02 (0.95-1.10) | 1.01 (0.93-1.09) | 1.04 (0.95-1.12) | 1.25 (1.14-1.36) | 1.04 (1.02-1.06) | <0.001 |
| Model 3 (n=78,110) | 1.05 (1.02-1.08) | 1 [reference] | 1.02 (0.95-1.10) | 1.03 (0.95-1.11) | 1.03 (0.95-1.12) | 1.19 (1.09-1.30) | 1.03 (1.01-1.06) | 0.001 |
| Model 4 (n=78,110) | 1.05 (1.02-1.08) | 1 [reference] | 1.02 (0.95-1.10) | 1.02 (0.95-1.11) | 1.03 (0.95-1.12) | 1.19 (1.09-1.29) | 1.03 (1.01-1.05) | 0.001 |
| ***Men*** |  |  |  |  |  |  |  |  |
| Model 1 (n=63,574) | 1.07 (1.05-1.10) | 1 [reference] | 0.99 (0.92-1.06) | 0.97 (0.90-1.04) | 1.10 (1.02-1.19) | 1.23 (1.14-1.35) | 1.05 (1.03-1.07) | <0.001 |
| Model 2 (n=63,440) | 1.06 (1.03-1.09) | 1 [reference] | 1.00 (0.93-1.07) | 0.97 (0.90-1.05) | 1.08 (1.00-1.17) | 1.18 (1.09-1.29) | 1.04 (1.02-1.06) | <0.001 |
| Model 3 (n=61,231) | 1.02 (1.00-1.05) | 1 [reference] | 1.00 (0.93-1.07) | 0.95 (0.89-1.03) | 1.03 (0.95-1.11) | 1.05 (0.97-1.15) | 1.01 (0.99-1.03) | 0.22 |
| Model 4 (n=61,230) | 1.02 (1.00-1.05) | 1 [reference] | 1.00 (0.93-1.07) | 0.96 (0.89-1.03) | 1.03 (0.95-1.12) | 1.05 (0.97-1.15) | 1.01 (0.99-1.03) | 0.2 |

*Note*: Data show ORs and 95% CIs unless otherwise indicated. P for trend is from regression models with quintiles. Model 1, unadjusted; model 2, adjusted for age, sex, and BMI (body mass index); model 3, model 2 additionally adjusted for smoking, alcohol, physical activity, ethnicity, and TDI (Townsend deprivation index at recruitment); model 4, model 3 additionally adjusted for diabetes and cardiovascular disease; *: CRP concentration was log transformed; Median CRP level was 1.15 mg/L (range 0.08-78.22 mg/L).

#### Test for specificity or commonality of association of CRP with depression and anxiety

Results for probit regression using probable diagnoses of depression and GAD as outcomes were similar to results from continuous symptom outcomes. The OR for probable depression (OR=1.029; 95% CI, 1.026-1.033) was larger than for probable GAD (OR=1.016; 95% CI, 1.012-1.020); *P-*value for difference <0.001. In logistic regression analyses, evidence for association of CRP with probable depression remained after adjusting for probable GAD (OR=1.09; 95% CI, 1.06-1.12), but not *vice versa* (OR=1.01; 95% CI, 0.97-1.04).

#### Results for Mendelian randomization analyses

##### Supplementary Table 4. Mendelian randomisation analysis of association of CRP and IL-6 with depression and anxiety using alternative genetic instruments

| **MR Approach** | **Exposure** | **Outcome** | **OR (95% CI)** | **P** |
| --- | --- | --- | --- | --- |
| **1-sample** |  |  |  |  |
| Inverse variance weighted | CRP (CCGC) | Depressive symptom score | 0.92 (0.79-1.08) | 0.318 |
| Inverse variance weighted | CRP (CCGC) | Anxiety symptom score | 0.90 (0.81-1.00) | 0.043 |
| Inverse variance weighted | CRP (CCGC) | Probable depression | 1.03 (0.90-1.19) | 0.669 |
| Inverse variance weighted | CRP (CCGC) | Probable GAD | 0.89 (0.76-1.05) | 0.159 |
| Inverse variance weighted | IL-6 (Swerdlow) | Depressive symptom score | 1.21 (0.96-1.52) | 0.114 |
| Inverse variance weighted | IL-6 (Swerdlow) | Anxiety symptom score | 1.16 (0.85-1.59) | 0.338 |
| Inverse variance weighted | IL-6 (Swerdlow) | Probable depression | 1.03 (0.77-1.37) | 0.855 |
| Inverse variance weighted | IL-6 (Swerdlow) | Probable GAD | 1.47 (1.06-2.04) | 0.021 |
| Wald Ratio | IL-6 (Sarwar) | Depressive symptom score | 1.17 (0.85-1.59) | 0.332 |
| Wald Ratio | IL-6 (Sarwar) | Anxiety symptom score | 1.07 (0.81-1.43) | 0.623 |
| Wald Ratio | IL-6 (Sarwar) | Probable depression | 1.05 (0.72-1.54) | 0.788 |
| Wald Ratio | IL-6 (Sarwar) | Probable GAD | 1.32 (0.86-2.04) | 0.203 |
| **2-sample** |  |  |  |  |
| Inverse variance weighted | CRP (CCGC) | Depressive symptom score | 0.88 (0.73-1.07) | 0.213 |
| Inverse variance weighted | CRP (CCGC) | Anxiety symptom score | 0.88 (0.78-1.00) | 0.056 |
| Inverse variance weighted | CRP (CCGC) | Probable depression | 1.02 (0.86-1.21) | 0.796 |
| Inverse variance weighted | CRP (CCGC) | Probable GAD | 0.85 (0.70-1.03) | 0.104 |
| Inverse variance weighted | IL-6 (Swerdlow) | Depressive symptom score | 0.84 (0.66-1.06) | 0.137 |
| Inverse variance weighted | IL-6 (Swerdlow) | Anxiety symptom score | 0.87 (0.63-1.21) | 0.408 |
| Inverse variance weighted | IL-6 (Swerdlow) | Probable depression | 0.99 (0.74-1.32) | 0.953 |
| Inverse variance weighted | IL-6 (Swerdlow) | Probable GAD | 0.69 (0.48-0.99) | 0.044 |
| Wald Ratio | IL-6 (Sarwar) | Depressive symptom score | 0.95 (0.86-1.05) | 0.332 |
| Wald Ratio | IL-6 (Sarwar) | Anxiety symptom score | 0.98 (0.89-1.07) | 0.623 |
| Wald Ratio | IL-6 (Sarwar) | Probable depression | 0.98 (0.87-1.12) | 0.788 |
| Wald Ratio | IL-6 (Sarwar) | Probable GAD | 0.91 (0.79-1.05) | 0.203 |

*Note*: 2-sample MR estimates for IL-6 instruments based on Swerdlow *et al.*^13^ and Sarwar *et al.*^14^ are expected to show reverse directionality as compared to the Georgakis *et al.*^12^ instrument for IL-6 and 1-sample instruments for IL-6 as the latter are all weighted on CRP concentrations. Genetic associations with IL-6 concentrations have been previously shown to be associated with reduced IL-6 signalling as key SNPs included in the instruments (rs2228145 [Sarwar] & rs7529229 [Swerdlow], that are in high LD in European populations^25^, R^2^=0.94) are also associated with increased sIL-6R levels that, together with gp130, provide a buffer against IL-6 activity under physiological conditions.^16^

##### Supplementary Table 5. IVW Mendelian randomisation analysis of association of IL-6 and CRP with depression and anxiety using Inverse Probability Weighted Outcome Associations

|  | **Depression Symptom Score** | | **Probable depression** | | **Anxiety Symptom Score** | | **Probable GAD** | |
| --- | --- | --- | --- | --- | --- | --- | --- | --- |
| **Model** | **OR (95% CI)** | ***P*-value** | **OR (95% CI)** | ***P*-value** | **OR (95% CI)** | ***P*-value** | **OR (95% CI)** | ***P*-value** |
| ***CRP*** |  |  |  |  |  |  |  |  |
| 2-Sample MR | 0.89 (0.79-0.99) | 0.04 | 0.95 (0.85-1.07) | 0.409 | 0.89 (0.81-0.97) | 0.008 | 0.86 (0.75-0.98) | 0.025 |
| 1-Sample MR | 0.90 (0.79-1.02) | 0.107 | 1.00 (0.88-1.14) | 0.98 | 0.89 (0.81-0.99) | 0.024 | 0.87 (0.75-1.02) | 0.096 |
| ***IL-6*** |  |  |  |  |  |  |  |  |
| 2-Sample MR | 1.30 (1.01-1.66) | 0.041 | 1.06 (0.8-1.42) | 0.673 | 1.07 (0.86-1.34) | 0.529 | 1.11 (0.8-1.54) | 0.535 |
| 1-Sample MR | 1.28 (1.01-1.62) | 0.039 | 1.10 (0.84-1.45) | 0.487 | 1.05 (0.86-1.30) | 0.619 | 1.04 (0.76-1.42) | 0.812 |

##### Supplementary Table 6. Heterogeneity statistics for IVW Mendelian randomisation analysis of association of IL-6 and CRP with depression and anxiety

| **Exposure** | **Outcome** | **Cochran’s Q** | **P** | **DF** |
| --- | --- | --- | --- | --- |
| ***1-sample MR*** |  |  |  |  |
| CRP (CCGC) | Depressive symptom score | 5.46 | 0.141 | 3 |
| CRP (CCGC) | Anxiety symptom score | 1.17 | 0.759 | 3 |
| CRP (CCGC) | Probable depression | 2.08 | 0.557 | 3 |
| CRP (CCGC) | Probable GAD | 3.05 | 0.384 | 3 |
| CRP (Georgakis) | Depressive symptom score | 29.7 | 0.158 | 23 |
| CRP (Georgakis) | Anxiety symptom score | 16.09 | 0.852 | 23 |
| CRP (Georgakis) | Probable depression | 20.57 | 0.608 | 23 |
| CRP (Georgakis) | Probable GAD | 23.87 | 0.411 | 23 |
| IL-6 (Georgakis) | Depressive symptom score | 6.78 | 0.342 | 6 |
| IL-6 (Georgakis) | Anxiety symptom score | 2.62 | 0.855 | 6 |
| IL-6 (Georgakis) | Probable depression | 2.57 | 0.861 | 6 |
| IL-6 (Georgakis) | Probable GAD | 2.37 | 0.883 | 6 |
| IL-6 (Swerdlow) | Depressive symptom score | 1.38 | 0.502 | 2 |
| IL-6 (Swerdlow) | Anxiety symptom score | 4.2 | 0.123 | 2 |
| IL-6 (Swerdlow) | Probable depression | 1.13 | 0.569 | 2 |
| IL-6 (Swerdlow) | Probable GAD | 2.05 | 0.359 | 2 |
| **2-sample MR** |  |  |  |  |
| CRP (CCGC) | Depressive symptom score | 4.02 | 0.134 | 2 |
| CRP (CCGC) | Anxiety symptom score | 1.06 | 0.589 | 2 |
| CRP (CCGC) | Probable depression | 2.05 | 0.359 | 2 |
| CRP (CCGC) | Probable GAD | 2.18 | 0.336 | 2 |
| CRP (Georgakis) | Depressive symptom score | 27.9 | 0.219 | 23 |
| CRP (Georgakis) | Anxiety symptom score | 13.92 | 0.929 | 23 |
| CRP (Georgakis) | Probable depression | 19.93 | 0.646 | 23 |
| CRP (Georgakis) | Probable GAD | 20.47 | 0.613 | 23 |
| IL-6 (Georgakis) | Depressive symptom score | 6.48 | 0.372 | 6 |
| IL-6 (Georgakis) | Anxiety symptom score | 2.41 | 0.878 | 6 |
| IL-6 (Georgakis) | Probable depression | 3 | 0.808 | 6 |
| IL-6 (Georgakis) | Probable GAD | 1.77 | 0.939 | 6 |
| IL-6 (Swerdlow) | Depressive symptom score | 1.68 | 0.433 | 2 |
| IL-6 (Swerdlow) | Anxiety symptom score | 4.56 | 0.102 | 2 |
| IL-6 (Swerdlow) | Probable depression | 1.16 | 0.560 | 2 |
| IL-6 (Swerdlow) | Probable GAD | 2.48 | 0.289 | 2 |

### SUPPLEMENTARY DISCUSSION

#### Potential interpretations for divergent effects of CRP and IL-6

##### Supplementary Figure 3. Potential divergent effects of specific IL-6 signalling pathways on depression risk

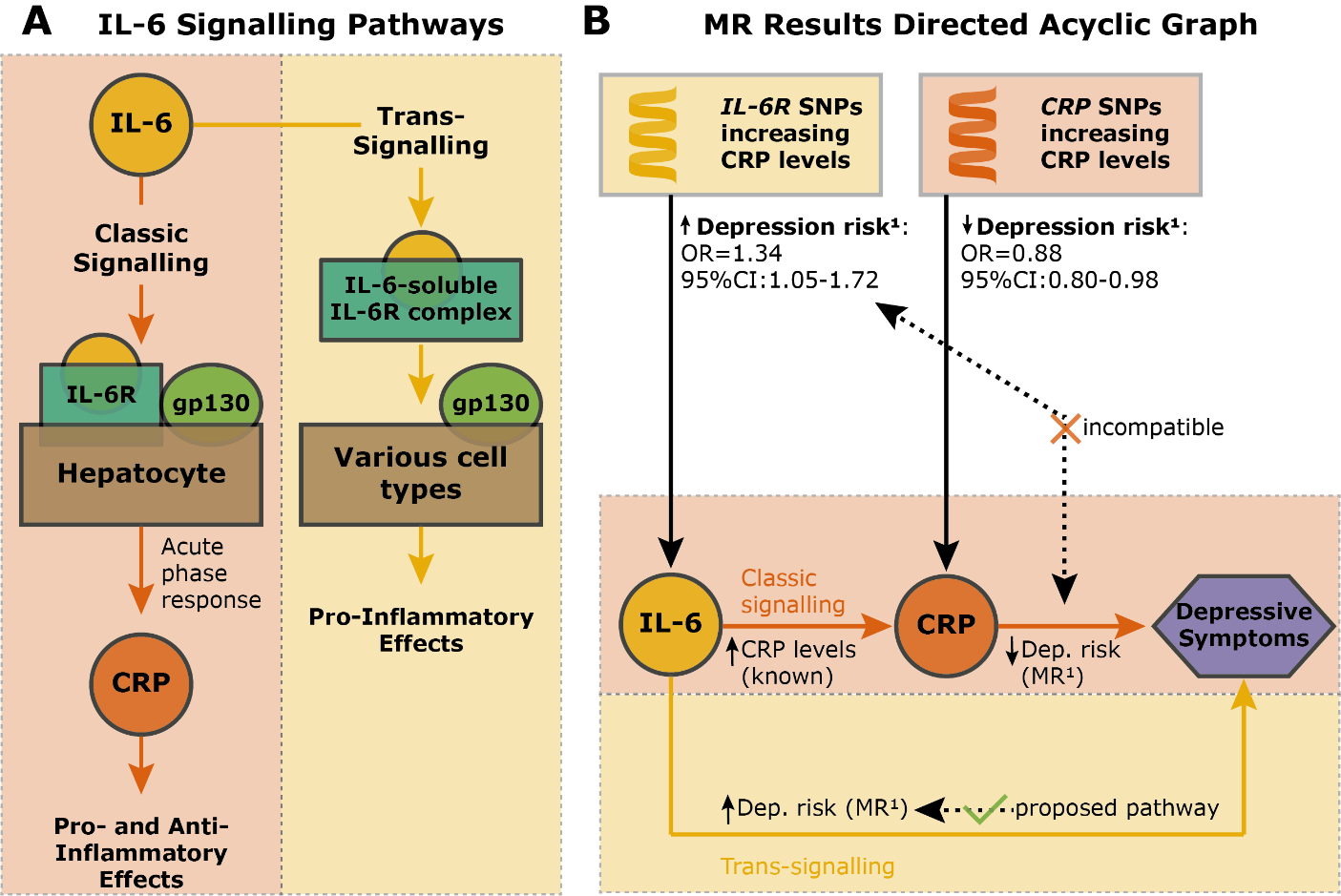

*Note*: Figure 1A shows IL-6 classic and trans-signalling pathways; see review by Hunter and Jones^26^. Figure 1B displays our working hypothesis arising from MR results that IL-6 trans-signalling confers increased risk for depression. ^1^MR estimates are based on 2-sample MR analysis using Georgakis *et al.*^12^ genetic instruments and continuous depressive symptoms as outcome (cf. Table 4). Abbreviations: gp130=glycoprotein 130; Dep.=depression; CRP=C-reactive protein; IL-6=interleukin-6.
